# Analytical performance of the VITA™ point-of-care CD4 enumeration assay

**DOI:** 10.64898/2025.12.16.25342340

**Authors:** Chui Mei Ong, Aquiles R. Henriquez-Trujillo, Melissa Alamillo, Wei Huang, Ed Goldberg, Dorien Van den Bossche, Tinne Gils, Alan H.B. Wu

## Abstract

CD4+ T-cell enumeration remains critical for management of people with HIV (PWH). In resource-limited settings, point-of-care CD4 testing can increase accessibility and decrease turnaround time compared to flow cytometry. This study evaluated the performance of VITA™ point-of-care CD4 (Accesso Biotech; VITA™ CD4) compared to AQUIOS™ (Beckman Coulter; AQUIOS™) flow cytometry.

Remnant venous blood samples from PWH with known CD4 counts on FDA-cleared AQUIOS™ flow cytometry were tested twice with VITA™ CD4. Statistical analyses included descriptive statistics, correlation analysis, Bland-Altman agreement analysis, and Passing-Bablok regression. The coefficient of variation was calculated on duplicate measurements. The University of San Francisco Institutional Review Board approved the study.

VITA^TM^ CD4 results showed strong correlation with AQUIOS flow cytometry (r = 0.973, p < 0.001). Bland-Altman analysis revealed a mean bias of -48.9 cells/µl (95% CI: -61.8 to -36.0), with limits of agreement from -231.1 to 133.3 cells/µl. For samples <500 CD4 cells/µL (n=137), the mean bias was reduced to -8.6 cells/µl (95% CI: -17.8 to 0.6). Passing-Bablok regression suggested a small constant positive bias and proportional bias at counts above 500 cells/µl [y = 30.2 (95% CI: 19.3 to 44.2) + 0.83x (95% CI: 0.80-0.86)]. The coefficient of variation for duplicate VITA POC CD4 measurements was 11.86%.

VITA™ CD4 results correlated strongly with flow cytometry at clinically relevant CD4 values. Its diagnostic performance and applicability at point-of-care make it a promising CD4 tool, also in settings with limited laboratory infrastructure. Further clinical validation and assessment of user acceptability is recommended.

## Introduction

Around 630,000 people died of AIDS in 2024, with mortality largely driven by advanced HIV disease (AHD) (1). One in three people with HIV (PWH) presented or re-engaged with care with AHD globally (2). In 13 nationally representative household surveys in sub Saharan Africa, one in ten PWH had AHD (3). Considering the global funding crisis for HIV, excess deaths are likely to occur in the coming years due to AHD (4).

A CD4 count is an essential diagnostic tool to identify those with AHD, defined in adults/adolescent PWH as having a CD4≤200 cells/µl, a World Health Organization (WHO) stage 3 or 4 disease (5, 6). Clinical staging is not very sensitive for AHD, and more than half of AHD cases require a CD4 cell count for diagnosis (7, 8). Depending on the context and clinical presentation, CD4 results will trigger TB lipoarabinomannan (TB-LAM) and cryptococcal antigen screening to detect tuberculosis and cryptococcal meningitis; the most important causes of mortality from HIV (9). Furthermore, CD4 testing is necessary to define eligibility for cotrimoxazole. These diagnostic and treatment interventions have been recommended by the WHO for use at health facilities and in outreach since 2017 (5). However, implementation is poor in facilities and absent in communities, in part due to the lack of appropriate point-of-care CD4 tests (10, 11).

Following scale up of universal Test and Treat, and viral load testing for monitoring of antiretroviral treatment (ART) response, most countries in sub Saharan Africa shifted limited funding for CD4 testing in favor of viral load devices (12). The reduction in global demand for CD4 testing led two leading manufacturers of CD4 devices used in the Global South, including the point-of-care Abbott PIMA™ test (Abbott, Chicago, IL, USA; PIMA), to discontinue production, leaving a gap in access to CD4 testing (11). The point-of-care lateral flow device VISITECT™ CD4 Advanced Disease (AccuBio, Alva, UK; VISITECT) is available but is limited by its varying diagnostic performance, user challenges, and the binary nature of its results (i.e. above or equal to/below 200 cells/µl) (11, 13, 14).

The VITA™ Point-of-Care CD4 Enumeration Assay (Accesso Biotech, San Francisco, CA, USA; VITA™ CD4) is a novel point-of-care portable CD4 device, based on immunofluorescence imaging cytometry, providing a numeric CD4 count within 35 minutes (15). The assay has not yet been evaluated in clinical samples. We aimed to evaluate of diagnostic performance VITA™ CD4 on clinical samples, in comparison with the WHO prequalified AQUIOS™ CL Flow Cytometer (Beckman Coulter, Brea, CA, USA; AQUIOS™) (16).

## Materials and methods

### Design, population, period

This was a laboratory-based diagnostic accuracy study performed on anonymised leftover fresh whole blood samples collected between December 2024 and February 2025, from adult (≥18 years old) PWH who attended the General Hospital of San Francisco. Samples were preselected to overrepresent lower CD4 ranges (>65% with CD4 ≤500 cells/µl), as this was the last threshold used for treatment initiation prior to introduction of universal Test and Treat (17). Data were accessed for analysis on 03/06/2025.

### VITA™ CD4 assay principle

An illustration of the assay principle is shown in fig 1. The whole blood sample is stained with fluorescence-labeled anti-CD3 and anti-CD4 antibodies in the dried reagent tube. The sample is then loaded into the cartridge and flows into the fluidic channel where the blood cells are presented as a thin layer. After inserting the cartridge into the VITA™ CD4 analyzer, cells are analysed by fluorescence microscopy. Specifically, a LED illumination is turned on corresponding to the excitation wavelength of a fluorophore-labeled antibody, and the specific emission filter is positioned in front of a complementary metal-oxide semiconductor sensor for the digital camera. Following the first fluorescence image, a different LED and emission filter is engaged to take the fluorescence image corresponding to the second antibody. Image processing and cell analysis algorithms are performed in the on-board computer and finally absolute CD4+ T cell count is reported based on the sampling volume calculated from the channel height and image area.

**Fig 1:**
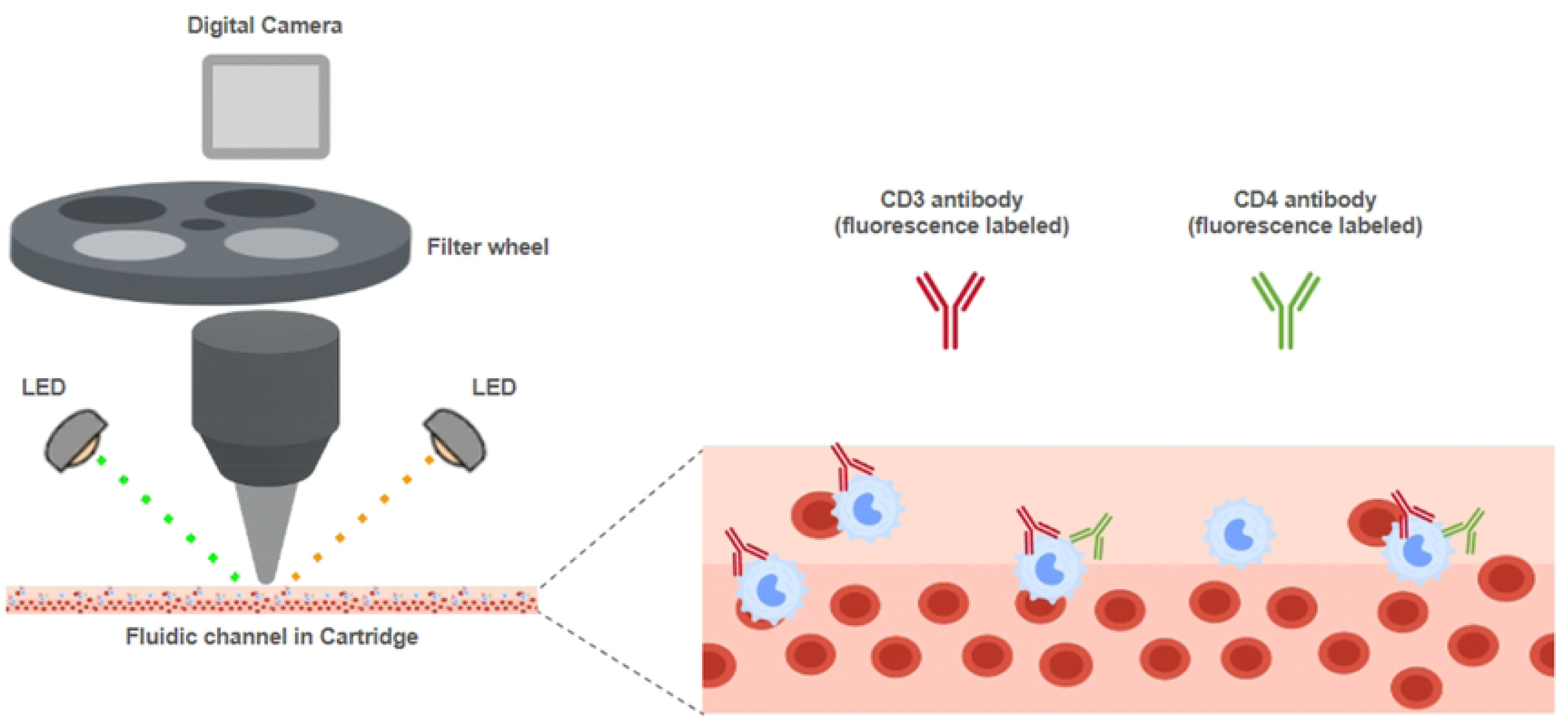
VITA™ CD4 assay principle.

### Procedure

The VITA™ CD4 test kit includes a pouch with VITA™ CD4 test cartridge and a dried reagent tube, and a sealing sticker. The VITA™ CD4 test procedure was performed by a laboratory technician as follows: The cartridge and the dried reagent tube were removed from the pouch. Remnant whole blood samples in EDTA tubes were aliquoted in a cryovial tubes and tested within 48h after phlebotomy. The specimen was thoroughly mixed by slowly inverting the cryovial tube 15 times with the cap firmly secured. Approximately 20 µl of whole blood specimen was aspirated with a volumetric pipette and the entire volume dispensed into the dried reagent tube. The sample was slowly pipetted up and down 20 times to rehydrate the reagent and mix with the blood. After the last mixing cycle, the mixture was carefully transferred into the loading port of the cartridge. The operator visually confirmed that the blood sample flowed through the cartridge chamber and reached the opening window at the end of the chamber, then placed the sealing sticker over the loading port so that the entire loading port was covered. The cartridge was then placed to incubate protected from light exposure (e.g. in a drawer) at room temperature for 30 minutes. Subsequently, the cartridge was inserted into the VITA™ CD4 analyzer. Following the on-screen instructions, sample information was entered on the touch screen and the analysis initiated. After 1-2 minutes, the numeric CD4 result was reported on the device screen. In case of an anomaly, a quality control error is reported instead of a CD4 result. All samples were analysed in duplicate with two cartridges on the same VITA™ CD4 analyser, with the duplicate analysis ran immediately after the first one. The AQUIOS™ procedure was performed per manufacturer instructions (18).

### Analysis

In case of error with the first VITA™ CD4 result, the duplicate result was used for analysis. Statistical analyses were performed using MedCalc® Statistical Software version 23.3.7 (MedCalc Software Ltd, Ostend, Belgium). Descriptive statistics were calculated for both methods. The VITA™ CD4 error rate, was calculated as the average proportion of erroneous results between the duplicates. The coefficient of variation (CV) was calculated from duplicate measurements with VITA^TM^ CD4. Method comparison analyses included Pearson correlation coefficient to assess the linear relationship between methods, Bland-Altman analysis to evaluate agreement and identify systematic bias, Passing-Bablok regression to assess constant and proportional differences. Subgroup analysis was performed for samples with CD4 counts <500 cells/µL, representing the clinically relevant range for treatment decisions (5).

Classification of VITA^TM^ CD4 at decision thresholds (200 cells/µl (corresponding to the definition of AHD (5)), and 500 cells/µl) was assessed by applying the Passing-Bablok regression equation to predict VITA^TM^ CD4 values, with 95% confidence intervals calculated using bootstrap resampling. Sensitivity and specificity to detect CD4 counts <200 cells/µl of VITA^TM^ CD4 compared to AQUIOS™, was presented with 95% confidence intervals. Statistical significance was set at p<0.05.

### Ethics statement

The study received ethical approval from the Institutional Review Board of the University of California San Francisco (Reference: 24-41393). No consent was required as the analysis was performed on anonymized leftover samples.

## Results

### Sample characteristics

A total of 203 samples were tested with AQUIOS™ and twice with VITA™ CD4 (test 1 and 2) (S1 VITA dataset). Seven errors occurred on test 1 (3.4%), five on test 2 (2.5%). The average error rate was 2.9% (12/406). Two samples with errors on both tests were excluded from further analyses. Two-hundred and one samples were included for the primary outcome analysis; consisting of 196 test results 1, and five test 2 results (Table 1).

**Table 1:**
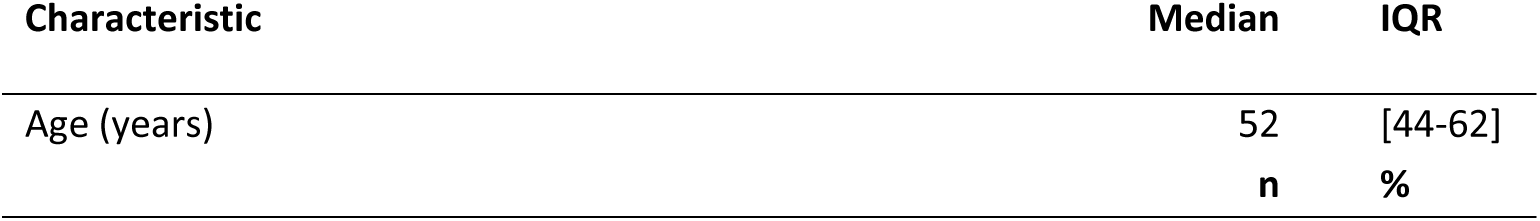
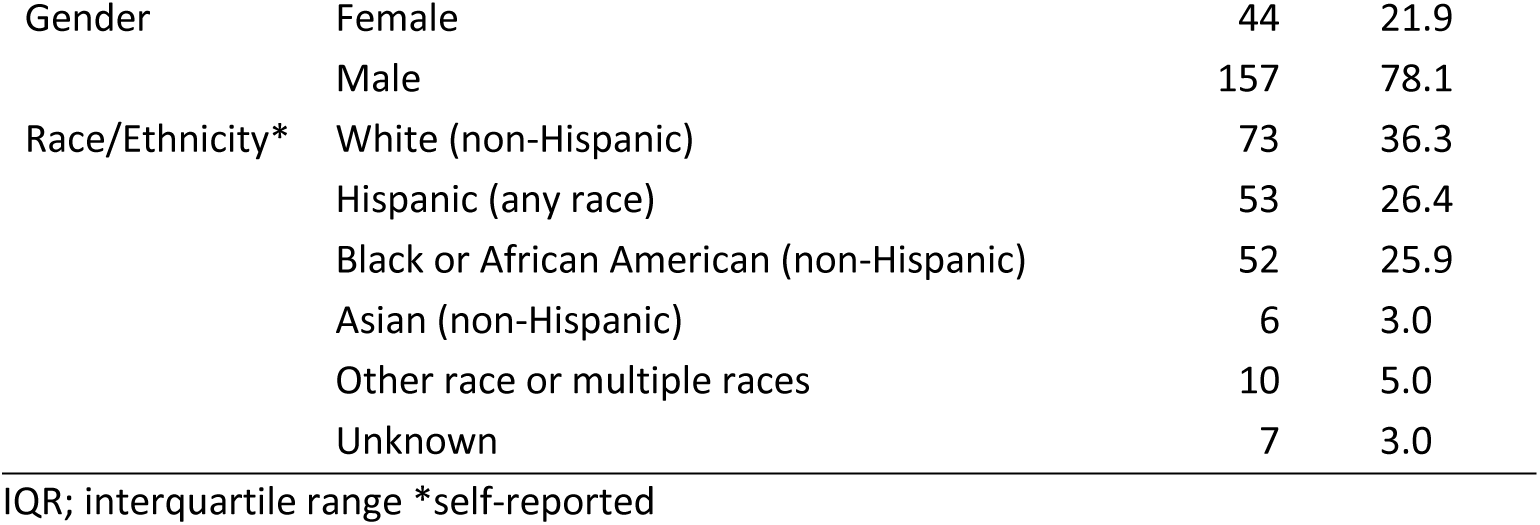
Demographics of participants with samples included in primary analysis (N=201)

| Characteristic | Median | IQR |
| --- | --- | --- |
| Age (years) | 52 | [44-62] |
|  | n | % |

|  |  |  |  |
| --- | --- | --- | --- |
| Gender | Female | 44 | 21.9 |
|  | Male | 157 | 78.1 |
| Race/Ethnicity* | White (non-Hispanic) | 73 | 36.3 |
|  | Hispanic (any race) | 53 | 26.4 |
|  | Black or African American (non-Hispanic) | 52 | 25.9 |
|  | Asian (non-Hispanic) | 6 | 3.0 |
|  | Other race or multiple races | 10 | 5.0 |
|  | Unknown | 7 | 3.0 |

IQR; interquartile range \*self-reported

Among 201 samples, 137 (68.2%) had a reference CD4 <500 cells/µl. CD4 counts ranged from 14 to 1,413 cells/µl on VITA™ CD4 and 17 to 1,698 cells/µL on AQUIOS™. The mean (±SD) CD4 count was 391.7 ± 283.6 cells/µl for VITA™ CD4 and 438.0 ± 341.7 cells/µL for AQUIOS™. The median values were 311 and 326 cells/µl. (Fig 2).

**Fig 2:**
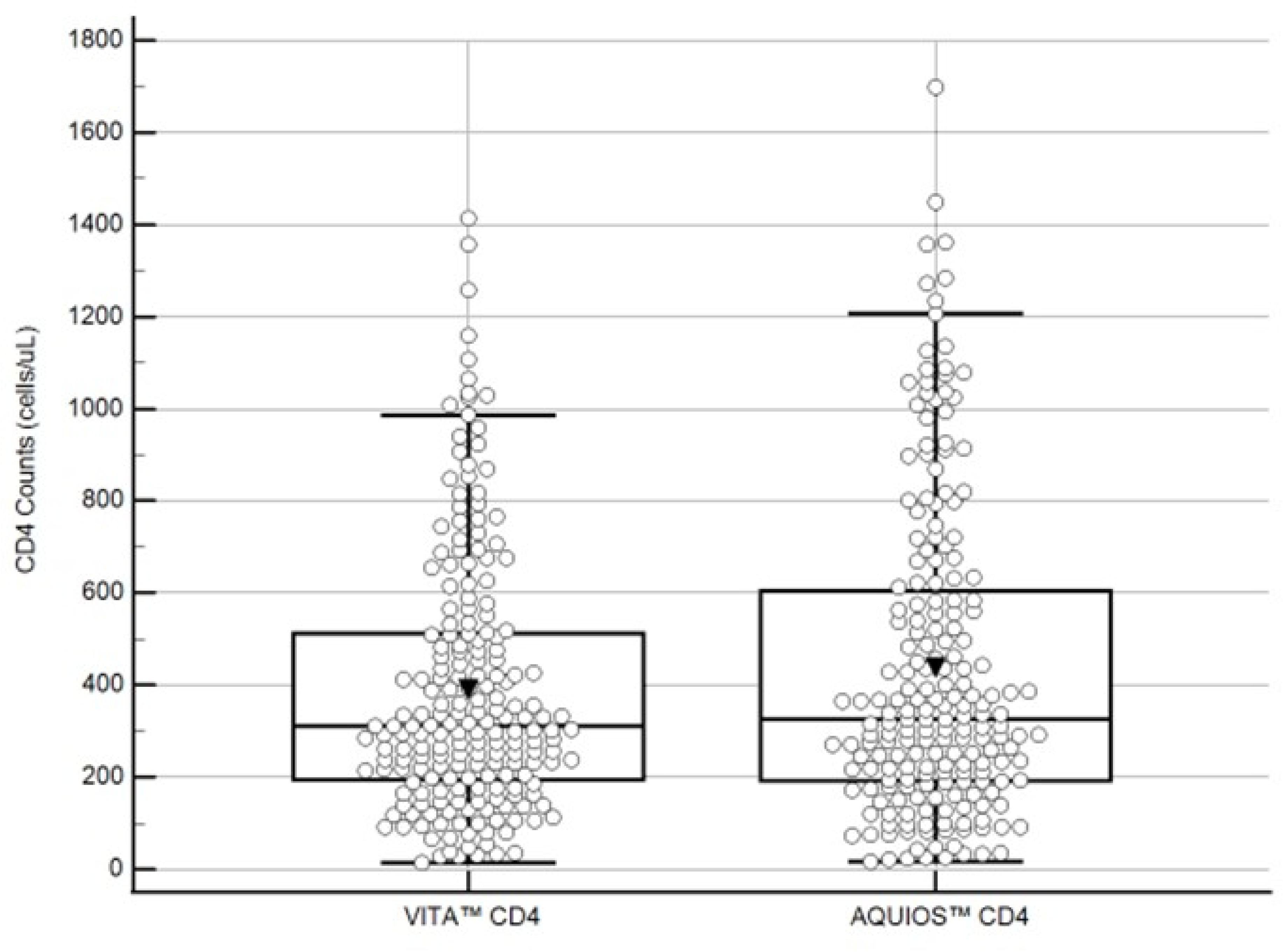
Boxplots for CD4 count distribution comparison between VITA™ CD4 and AQUIOS™ flow cytometry. N= 201.

Duplicate measurements were available for 193 samples. The within-subject standard deviation was 47.2 cells/µL, yielding a coefficient of variation of 11.86%.

### Correlation analysis

Strong positive correlation was observed between VITA and AQUIOS measurements (r = 0.973, 95% CI: 0.965-0.980, p < 0.001) across the entire measurement range.

### Bland-Altman Agreement analysis

Bland-Altman analysis revealed a mean bias of -48.9 cells/µl (95% CI: -61.8 to -36.0). The 95% limits of agreement were -231.1 to 133.3 cells/µl (Fig 3).

**Fig 3:**
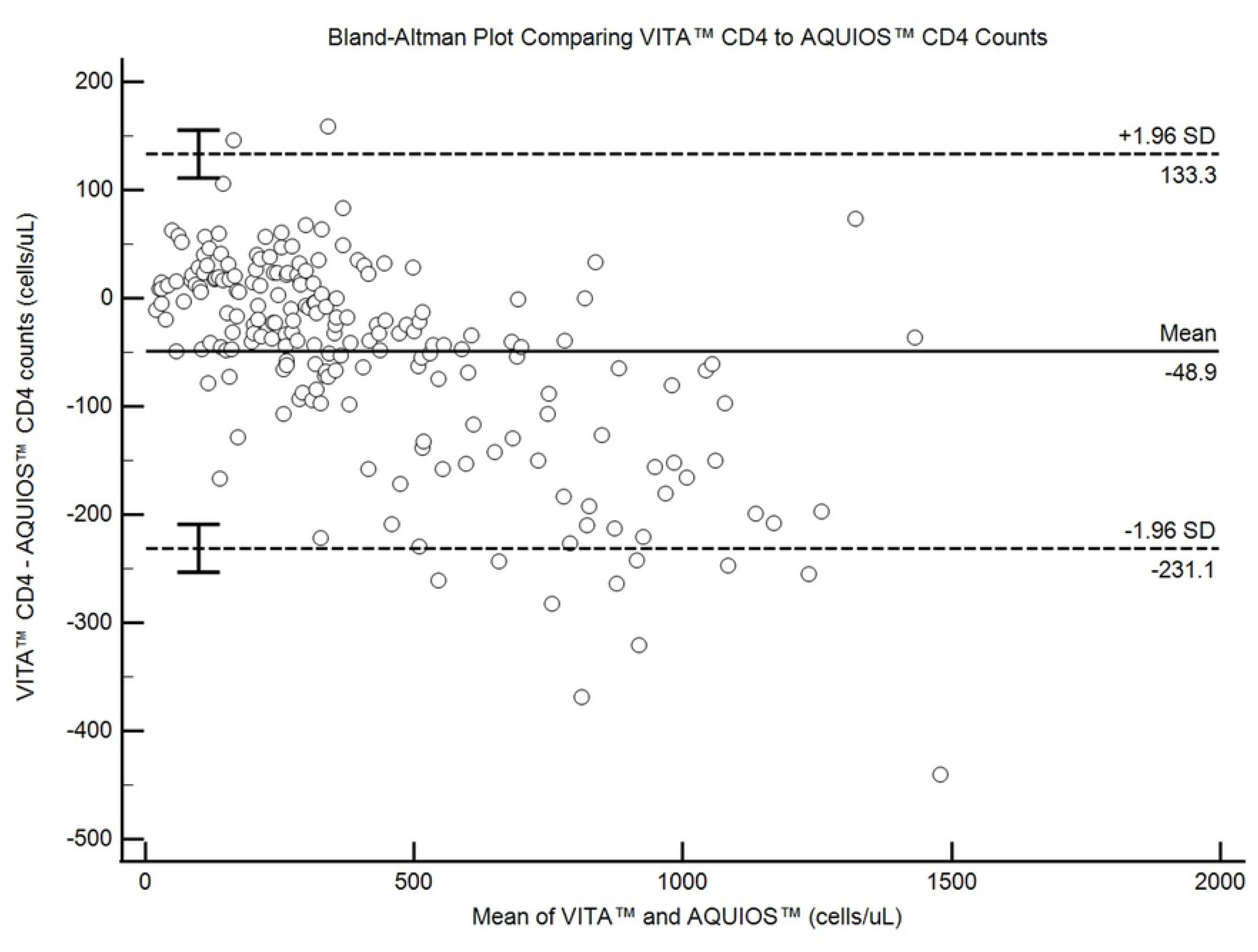
Bland-Altman plot for VITA™ CD4 versus AQUIOS™ flow cytometry with mean differences and limits of agreement.

For samples with CD4 counts <500 cells/µl (n=137), the mean bias was reduced to -8.6 cells/µl (95% CI: -17.8 to 0.6), with limits of agreement from -115.1 to 97.9 cells/µl (Fig 4).

**Fig 4:**
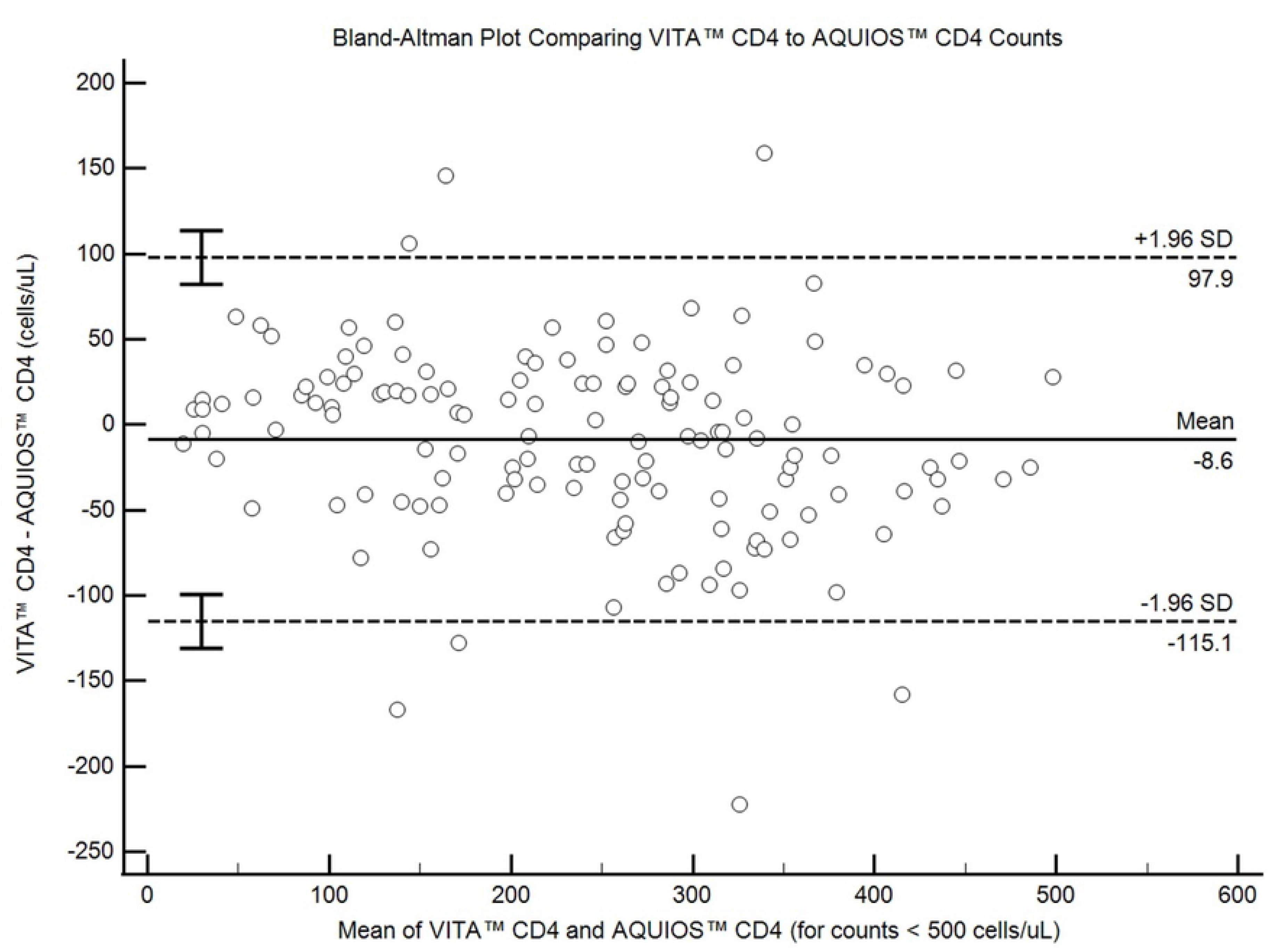
Bland-Altman plot for VITA™ CD4 versus AQUIOS™ flow cytometry with mean differences and limits of agreement for AQUIOS™ CD4 counts of <500 cells/µl.

### Regression analysis

Passing-Bablok regression analysis yielded the equation: VITA™ CD4 = 30.2 + 0.83 × AQUIOS™. The intercept of 30.2 (95% CI: 19.3-44.2) suggests a small constant positive bias, while the slope of 0.83 (95% CI: 0.80-0.86) indicates a proportional negative bias, with VITA™ CD4 reading approximately 17% lower than AQUIOS™ at higher CD4 counts (Fig 5).

**Fig 5.**
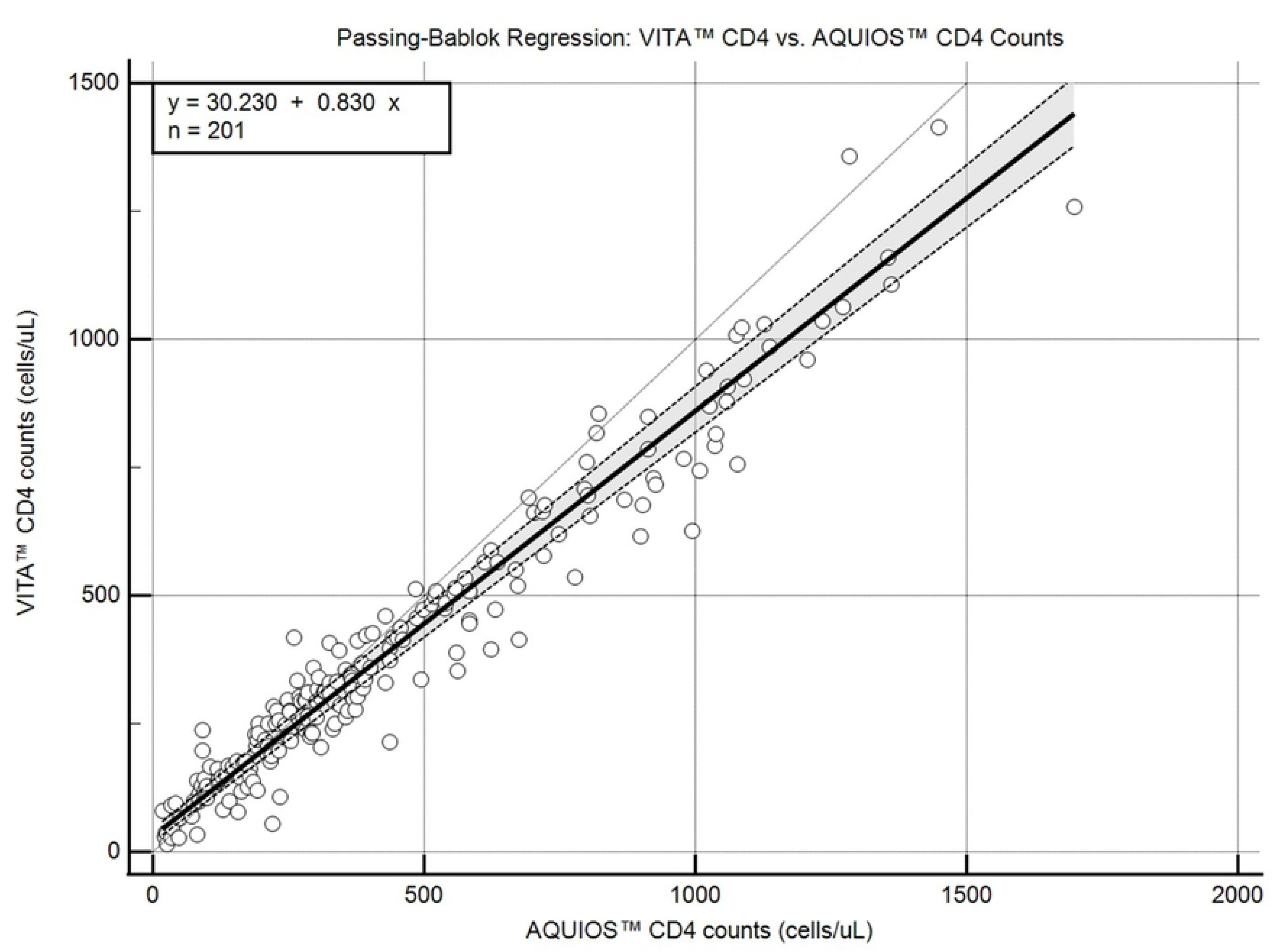
Passing-Bablok regression plot for VITA™ CD4 versus AQUIOS™ flow cytometry.

### Classification at Decision Thresholds

At the clinically important threshold of 200 cells/µL, the predicted VITA™ CD4 value was 196.3 cells/µL (95% CI: 188.6-206.3), representing a bias of -3.7 cells/µL (-1.84%). At 500 cells/µL, the predicted VITA™ CD4 value was 445.4 cells/µL (95% CI: 437.6-459.8), with a bias of -54.6 cells/µL (-10.91%). Sensitivity and specificity of VITA™ CD4 to detect a CD4 <200 cells/μl were 89% [95%CI: 77─96%] and 95% [95%CI: 91─98%], respectively.

## Discussion

This study demonstrates that CD4 results measured by VITA™ CD4 and AQUIOS™ correlated strongly. While a systematic negative bias of VITA™ CD4 was observed, the overall agreement between methods was excellent at CD4 counts below 500 cells/µL, where clinical decisions are most critical.

The observed negative bias of approximately 49 cells/µl in Bland-Altman analysis revealed that VITA™ CD4 systematically underestimated CD4 counts compared to AQUIOS™. This is consistent with findings from other point-of-care CD4 devices, which often show systematic differences compared to flow cytometry (19). The reduced bias at CD4 counts <500 cells/µl suggests that VITA performs best in the range where accurate enumeration is required to identify and manage patients with AHD, or start and stop cotrimoxazole prophylaxis (5).

The error rate of VITA™ CD4 was optimal (2.9%) when compared to the minimum of <5% described in a WHO target product profile for point-of-care tests to identify AHD. The level of precision of 11.86% is comparable to other point-of-care CD4 devices, and also above the minimal WHO threshold (<15%) (20).

There was a high concordance between VITA™ CD4 and AQUIOS™ results, with a high correlation coefficient (r= 0.973). The proportional bias identified through regression analysis (slope = 0.83) also indicates that differences between methods increase at higher CD4 counts, where no clinically relevant decision thresholds are situated. The minimal bias at the 200 cells/µL threshold (-1.84%) reassuring, as this represents a key decision point for reflex testing for opportunistic infections. VITA™ CD4 specificity to identify AHD was optimal (95%) and its sensitivity (89%) surpassed the minimal WHO threshold (WHO: minimal >80%, optimal >90%) (20). This highlights the potential for the VITA™ CD4 test to be used as part of the AHD care package by lay workers, who may rely on this threshold to perform additional TB-LAM or cryptococcal antigen testing at point-of-care without interpreting exact CD4 values. The recorded numeric count can be used for further clinical evaluation by medically trained staff.

Strengths of this study include the large sample size, comprehensive statistical analysis using multiple complementary approaches, and evaluation of precision through duplicate testing. The inclusion of samples across a wide CD4 range enhances generalizability. Limitations include the lack of clinical outcome data and absence of information on specific patient populations (e.g. samples with abnormal lymphocyte subsets). Additionally, the study was conducted in a controlled laboratory setting, and performance in field conditions requires further evaluation.

User acceptance and VITA™ CD4 usability should be assessed. Accuracy of VITA™ CD4 when performed on finger prick samples should be evaluated in parallel to whole blood samples.

The WHO confirmed CD4 testing is the preferred way to assess AHD in its recent recommendations (6). A major gap exists in the availability of CD4 measurement tools, especially for use at point-of-care. Compared to flow cytometry, VITA^TM^CD4 device offers advantages for resource-limited settings, including rapid turnaround time, minimal infrastructure requirements, and simplified operation. The other available point-of-care CD4 test, VISITECT CD4 Advanced Disease, has multiple limitations, including a variable specificity, and variable acceptability for healthcare workers following its result depending on reader interpretation of a color intensity (13–15, 21–23). VITA™ CD4 has the advantage of providing a numeric CD4 count, necessary to trigger different clinical interventions. Due to the market scarcity for CD4 tests, complementary roles could be taken by different CD4 tests. Cost-effectiveness and feasibility of different approaches should be assessed.

## Conclusions

The VITA^TM^ CD4 assay demonstrated strong correlation and clinically acceptable agreement with reference flow cytometry for CD4 enumeration. Although a systematic negative bias was observed, VITA^TM^ CD4 performance was excellent at clinically relevant CD4 levels. The assay’s precision, portability, rapid turnaround time, and ability to generate numeric CD4 counts support its use as a practical alternative to flow cytometry in settings requiring rapid, decentralized CD4 testing. Further evaluation under routine primary care conditions and assessment of user acceptability are warranted.

## Author contributions

CMO, ARHT, & AHBW designed the study. WH and EG developed and provided the VITA™ CD4 test. Funding was acquired by AHBW. The project was administered by CMO & MA. AHBW supervised the study. ARHT conducted formal analysis. TG and DvB wrote the first draft and all authors reviewed and approved the final manuscript.

## Data availability

Data supporting this study are included as supporting materials.

## Supporting information

S1 VITA dataset

